# Challenges in Implementing a Mobile AI Chatbot Intervention for Depression Among Youth on Psychiatric Waiting Lists: A Randomized Control Study Termination Report

**DOI:** 10.1101/2024.11.25.24317880

**Authors:** Junichi Fujita, Yuichiro Yano, Satoru Shinoda, Noriko Sho, Masaki Otsuki, Akira Suda, Mizuho Takayama, Tomoko Moroga, Hiroyuki Yamaguchi, Mio Ishii, Tomoyuki Miyazaki

## Abstract

**Background:** The mental health of children and adolescents is a growing public health concern, with increasing rates of depression and anxiety impacting their emotional, social, and academic well-being. In Japan, access to timely psychiatric care is limited, leading to extended waiting periods that can range from three months to a year. AI-driven chatbots, such as emol, which integrate Acceptance and Commitment Therapy (ACT), show potential as digital solutions to support young patients during these waiting times. The AI chatbot emol was selected based on a comprehensive review of Japanese mental health technology applications, including in-person evaluations with company representatives.

**Objective:** This exploratory, parallel-group randomized controlled trial examined the feasibility of using AI chatbot emol with pediatric and adolescent individuals on psychiatric waiting lists.

**Methods:** Participants aged 12-18 years were recruited from four hospitals in Kanagawa Prefecture and randomly assigned to either an intervention group, receiving eight weekly chatbot sessions, or a control group, receiving standard mental health information. The primary outcome was change in Patient Health Questionnaire-9 (PHQ-9) scores from pre- to post-intervention. Secondary assessments, such as voice and writing pressure analysis, provided additional engagement metrics, with data collected at baseline, during the intervention, and at week 9.

**Results:** Of the 96 eligible individuals on psychiatric waiting lists, eight expressed interest, and three provided initial consent. However, all participants subsequently withdrew or were excluded, resulting in no evaluable data for analysis. Low engagement may have been influenced by the perceived irrelevance of digital tools, complex protocols, and privacy concerns.

**Conclusions:** Significant barriers to engagement suggest that digital interventions may need simpler protocols and trusted environments to improve feasibility. Future studies could test these interventions in supportive settings, like schools or community centers, to enhance accessibility and participation among youth.

**Trial Registration:** Japan Registry of Clinical Trials (jRCT) jRCT1032230427

## INTRODUCTION

Depression and anxiety in children and adolescents are increasingly recognized as significant public health issues, profoundly affecting their emotional, social, and academic development [1,2]. Early intervention is crucial, as untreated depressive symptoms in youth are associated with a higher risk of recurrent episodes in adulthood [3]. Although traditional face-to-face mental health services have been the standard approach, research indicates that adolescents and young adults often prefer online mental health support over in-person consultations. A previous study found that young people’s preferences for mental health help-seeking vary significantly by age and stage of development, with adolescents showing greater inclination toward online resources due to increased privacy, reduced stigma, and enhanced autonomy [4]. This preference for digital mental health support is especially relevant for those with social anxiety, depression, or concerns about confidentiality. Digital interventions may provide a more accessible entry point to mental health care for youth who might otherwise avoid seeking traditional services, potentially addressing treatment gaps during critical developmental periods.

Despite the growing demand for effective treatments, access to child and adolescent psychiatric services remains limited due to resource shortages, resulting in extended waiting periods for consultation worldwide [5]. In Japan, access to child and adolescent mental health services remains a significant challenge, with a severe shortage of trained specialists and long waiting times for consultation. Previous studies have highlighted that the mental health system is primarily focused on adult and geriatric care, leaving pediatric mental health services underdeveloped and difficult to access. The limited availability of trained professionals contribute to prolonged waiting periods for care [6]. At Yokohama City University Hospital, for example, as of October 2023, the waiting time for appointments was 8 months, with over 120 patients aged 12-18 years on the psychiatric waiting list. The impact of the COVID-19 pandemic has further aggravated the mental health of young individuals globally, with rates of depression and anxiety among children and adolescents approximately doubling compared to pre-pandemic levels [7]. The increasing prevalence of adolescent depression and prolonged waiting periods for psychiatric care present urgent challenges in mental health service delivery. For young patients on waiting lists, the lack of timely intervention can exacerbate symptoms and increase the risk of severe outcomes, such as self-harm, suicide attempts, or psychotic experiences [8, 9].

Cognitive behavioral therapy (CBT), such as Acceptance and Commitment Therapy (ACT) is a first-line treatment for depression and anxiety in young people, yet barriers such as limited availability of trained therapists highlight the need for alternative delivery methods [10]. Digital health technologies, including AI-driven chatbots, offer an innovative approach to bridge the gap between service demand and supply. Existing studies have shown that internet-delivered CBT can be as effective as therapist-delivered CBT for treating depressive symptoms in children and adolescents [11]. Preliminary research on AI chatbots has demonstrated significant reductions in depression and anxiety among young users, underscoring the potential of these digital tools [12, 13].

Despite the growing interest in digital mental health interventions, there is limited research on the effectiveness of AI chatbots specifically for children and adolescents awaiting psychiatric consultation. Although prior studies have highlighted the potential benefits of AI chatbots in reducing depressive symptoms [14], these interventions have not been systematically tested among high-need populations, such as youth on psychiatric waiting lists. Given the prevalence of depressive symptoms among pediatric psychiatric patients and the potential for severe consequences, timely intervention is crucial [7, 8]. By offering mental health support through accessible digital devices like smartphones, the AI chatbot enables pre-consultation care that can be implemented in various settings, including schools. This approach could facilitate early intervention and support, providing a bridge until professional care becomes available and making mental health resources more accessible to young individuals outside traditional clinical environments.

Japan has a growing number of mental health technology applications, including various AI-driven chatbots, as outlined in recent market analyses. Several AI chatbots for mental health care have demonstrated reasonable feasibility, acceptability, and potential usefulness in Japan [15, 16]. However, despite the high smartphone adoption rate among young people (96.9% among those aged 18-29), actual utilization of health management services remains notably low at only 21.6% [17]. This gap between technology access and health-specific application usage highlights a significant opportunity for targeted digital mental health interventions. In the Japanese cultural context, healthcare services are evaluated through a balanced assessment where technical quality of care and healthcare staff behavior are both considered important factors that can compensate for each other in forming overall service quality judgments [18]. This tendency to value both technical expertise and interpersonal interactions may partially explain why Japanese patients prefer in-person medical consultations, potentially influencing the adoption rate of digital mental health interventions. Research on the social acceptance of smart health services in Japan has identified several key factors influencing adoption, including trust in service providers, perceived benefits and necessity, and risk perception regarding personal data protection [19]. These findings are particularly relevant for mental health applications where highly sensitive personal information is collected and utilized.

While various mental health chatbots exist in the Japanese market, few have been developed specifically for children and adolescents. For example, AI chatbot emol, developed in Japan, offers structured, ACT-based interventions through an accessible, engaging interface tailored for youth (Appendix). By leveraging its therapeutic design and user-friendly features, it provides interim support to adolescents awaiting professional care. Despite these technological advancements, there remains a significant research gap regarding the effectiveness of AI chatbots for supporting children and adolescents on psychiatric waiting lists. Given the extended waiting periods for psychiatric consultations in Japan and the increasing prevalence of mental health challenges among youth, investigating digital interventions that can provide interim support becomes particularly important. Previous studies have shown promising results for internet-delivered cognitive behavioral therapy among young people [11], but few have specifically examined AI-driven interventions for those awaiting professional psychiatric care.

We hypothesized that using the AI chatbot would significantly improve depressive symptoms and reduce clinical symptoms among children and adolescents on psychiatric waiting lists compared to standard care, thus enhancing mental health outcomes during the waiting period. The purpose of this study was to evaluate the effectiveness of the AI chatbot in improving depressive symptoms during the waiting period for children and adolescents on waiting lists, as a preliminary phase. The primary aims were to estimate the data acquisition rate and dropout rate in the intervention group, and to estimate the difference in depressive symptom change between the intervention and control groups before and after the intervention, to validate case number calculations for the next randomized controlled trial (RCT).

## METHODS

### Study Design

This exploratory, parallel-group, randomized controlled trial assigned participants to either an intervention group using the AI chatbot or a control group receiving general mental health information through a website publicly available, featuring clinical information that children and their families commonly review before their appointments. Unlike "standard care," which typically involves direct clinical assessment and treatment planning by a mental health professional, the control condition in this study only provided access to publicly available mental health education materials. No interactive therapeutic elements, personalized psychological interventions, or professional counseling were included in the control condition. This design reflects the real-world experience of many psychiatric waiting list patients in Japan, who often receive only basic mental health information while awaiting consultation.

Participants were centrally randomized by Nouvell Plus Inc. using a minimization method based on baseline PHQ-9 scores and gender to prevent significant imbalance. Blinding was not implemented as it was deemed unnecessary for the intervention.

Recruitment materials explicitly stated that the study was conducted by researchers from Yokohama City University. This information was included on printed leaflets distributed to potential participants. The affiliation was presented to enhance credibility but may also have influenced participant expectations and willingness to enroll. Participants were briefed about the study objectives, procedures, and potential risks through an online recruitment session. Written informed consent was obtained from both participants and guardians before enrollment. The informed consent documentation is available in the appendix for transparency.

No changes to the trial methods, including eligibility criteria, were made after trial commencement.

### AI Chatbot Selection Process

To identify the most suitable AI chatbot for supporting adolescents on psychiatric waiting lists, a comprehensive evaluation of existing mental health applications in Japan was conducted. The selection process involved the following steps:

#### 1. Comprehensive Review of Applications

Publicly available AI-driven mental health chatbots were reviewed for their therapeutic frameworks, usability, and relevance to adolescents. Special consideration was given to apps incorporating evidence-based treatments such as Acceptance and Commitment Therapy (ACT).

#### 2. In-Person Evaluation with Developers

Developers of shortlisted chatbots were interviewed to gain deeper insights into app features, target audiences, and implementation strategies.

#### 3. Selection Criteria

Chatbots were evaluated based on the following criteria:

・Integration of evidence-based therapeutic frameworks (e.g., ACT).

・Accessibility and ease of use for adolescents.

・Engagement-focused design, including gamified elements or interactive interfaces.

Compatibility with the study’s requirements for digital interventions in clinical settings.

Finally, the AI chatbot emol, developed by Emol Inc (Appendix). and released in March 2018, was selected for its integration of Acceptance and Commitment Therapy (ACT), an evidence-based treatment for depression and anxiety. The chatbot was developed in collaboration with clinical psychologists and researchers to integrate ACT based principles. The chatbot includes features such as conversational AI, emotional logging, interactive exercises, gamification elements, and monitoring tools to support user engagement. This therapeutic approach enables AI chatbot emol to offer targeted, structured interventions, distinguishing it from chatbots with less comprehensive therapeutic frameworks. The user interface employs a minimalist design, with simple text-based interactions and no external hyperlinks. Content was developed in collaboration with clinical psychologists and researchers at Emol Inc., ensuring alignment with ACT based principles. The chatbot provides asynchronous communication, allowing users to engage at their convenience without requiring real-time interaction.

AI chatbot emol’s design prioritizes accessibility and engagement, particularly for young users, by featuring a friendly AI character named "Roku." AI chatbot emol features a friendly character named "Roku," who guides users through ACT-based conversations in a relatable manner. The chatbot provides asynchronous communication, allowing participants to engage at their convenience. Sessions are pre-programmed to follow a consistent instructional strategy, beginning with a mood check-in and concluding with goal-setting exercises. Additional features, such as emotional logging and sleep tracking, allow users to actively monitor their mental health. AI chatbot emol’s comprehensive approach to digital mental self-care makes it especially well-suited for adolescents, who may prefer digital interactions over traditional therapy. By offering an approachable alternative for managing mental health challenges, AI chatbot emol has the potential to fill a critical gap in mental health service delivery for young people awaiting professional care.

However, no formal feasibility or usability studies have been conducted specifically for children and adolescents awaiting psychiatric consultation. The selection of emol for this study was based on a comprehensive review of Japanese mental health technology applications and direct discussions with the developers.

In this study, Emol Inc. provided the AI Chatbot emol at a discounted rate. This is disclosed in the Conflict of Interest section.

Details regarding the character design of AI Chatbot emol and its ACT-based interactions with the character "Roku" are provided in the Supplementary File. Additionally, the official website of Emol Inc., along with other relevant online resources, is listed in the Appendix for further reference.

### Participants

Participants in this study were individuals on the psychiatric waiting list for Yokohama City University Hospital, Yokohama City University Medical Center, Kanagawa Children’s Medical Center, and Fujisawa City Hospital, all four hospitals located in the south-eastern region of Kanagawa Prefecture. Those were aged between 12 and 18 years at the time of enrollment. These hospitals play a central role in child psychiatric services in the eastern part of Kanagawa Prefecture, Japan. As of January 2024, the population of 10-19-year-olds in Kanagawa Prefecture was estimated to be 774,283 (Appendix).

The recruitment period was planned to span from October 1st, 2023, to September 30th, 2024, allowing sufficient time to ensure robust participant enrollment and data collection.

### Inclusion and Exclusion Criteria

The inclusion criteria were as follows: 1) a score of 10 or higher on the Patient Health Questionnaire-9 (PHQ-9), indicating clinical depressive symptoms; 2) access to a device capable of running AI chatbot emol; 3) access to online interviews via Zoom (Zoom Inc.); 4) proficiency in reading and writing Japanese at the upper elementary school level; and 5) agreement to abstain from using other mental self-care apps during the study period.

The exclusion criteria were as follows: 1) patients deemed to require urgent care by a child psychiatrist; 2) patients reporting a suicide attempt within the past two weeks on their screening questionnaire, or patients in urgent need of physical treatment due to conditions such as anorexia; 3) patients who had received treatment at another psychiatric facility within the past month or who expressed a desire for future treatment at such a facility; 4) patients who were continuing to take psychotropic medications prescribed by the above-mentioned facilities; and 5) patients unable to complete diaries due to physical issues such as injury.

### Intervention Group

The intervention group received general mental health information via the Yokohama City University child psychiatry department’s website, “Oyako-no Kokoro-no Tomarigi” (Appendix), which provides short videos programs and texts that provide easy-to-understand explanations of common mental health issues for children and adolescents.

Additionally, eight weekly sessions were provided by the AI chatbot emol. Each session lasted between 20 to 30 minutes. No major bug fixes, system downtimes, content changes, or unexpected events occurred during the trial that could have influenced the intervention’s functionality or delivery. During the trial period, the version of emol remained unchanged, and no major content updates, feature modifications, or dynamic content changes occurred. The intervention was evaluated as a stable version, ensuring the replicability of study findings. While the current version reflects the intervention used in this study, future changes may occur. To ensure replicability, screenshots and videos of the interface are available upon request from the developers. Participants accessed the AI chatbot emol via their personal smartphones. All participants needed to have a stable internet connection and a device capable of running the application. Access to the chatbot was restricted to study participants only, and no public demo mode was available. However, a free sample version of the emol program, which differs from the study version, is publicly available at the website. Participants were not required to pay any fees for accessing the chatbot during the study. A stable internet connection and a compatible device were necessary for access. No critical secular events, such as changes in internet resources or hardware requirements, occurred during the study period.

While we await specific dialogue examples from Emol Inc. to further illustrate how these ACT processes were implemented conversationally, the session structure was designed to progressively build psychological flexibility through these interconnected processes. The detailed session structure and content are presented in Supplementary Table, and an example of user interaction with the AI character ’Roku’ is illustrated in Supplementary Figure.

The session overview in AI chatbot emol included the following:

1. Session 1 (Distress and suffering): Introduces psychological flexibility by helping users recognize their distress patterns, aligning with the Acceptance process of ACT.
2. Session 2 (Avoidance of experiences): Addresses Cognitive defusion by identifying experiential avoidance patterns and encouraging users to observe their thoughts rather than becoming entangled in them.
3. Session 3 (Control over what can be managed): Emphasizes Being Present and distinguishing between controllable and uncontrollable aspects of experience.
4. Session 4 (Acceptance): Deepens the Acceptance process by guiding users to embrace difficult emotions rather than struggling with them.
5. Session 5 (Observing from a detached perspective): Develops Self-as-Context by helping users adopt an observer perspective toward their experiences.
6. Session 6 (Life values): Explores the Values process by assisting users in identifying what matters most to them personally.
7. Session 7 (Commitment): Focuses on Committed Action by translating values into concrete behavioral goals.
8. Session 8 (Continuation of commitment): Reinforces Committed Action while integrating all ACT processes for sustainable change.

Weekly online assessments were conducted at Week 0, during the intervention period, and at Week 9. Weekly online assessments were conducted at Week 0, during the intervention period, and at Week 9. Non physician research assistants encouraged participants to use the pen consistently for their diary entries and performed minimal mental status checks during these assessments. No additional co-interventions, training sessions, or structured support were provided beyond these assessments. For routine application outside of the RCT setting, no training is required for users. No automated prompts or reminders were used during the trial or for routine application outside of the RCT setting. The periodic assessments were administered by non physician research assistants, who performed minimal mental status checks and checked assessment items such as PHQ-9, Athens Insomnia Scale (AIS), and adverse events. Simultaneously, non-physician research assistants saved recorded audio data for voice analysis. They also provided technical assistance when required. Human support was limited to assessment and monitoring during the intervention, with no direct involvement in the therapeutic process. For routine application outside of the RCT setting, no human involvement would be required. For measuring writing pressure, participants were provided with an intelligent pen pressure device developed by ZEBRA HOLDINGS Inc. and given a diary in which they recorded the date, time, weather, and mood (options included "good," "normal," or "bad") within two hours of waking. Research assistants encouraged participants to use the pen consistently for their diary entries.

### Control Group

The control group received general mental health information via the Yokohama City University child psychiatry department’s website, "Oyako-no Kokoro-no Tomarigi" (Appendix). This website provides educational resources about common mental health conditions in children and adolescents through easy-to-understand videos and text explanations specifically designed for young people. The video content features conversations between teddy bear and rabbit avatars discussing common mental health symptoms and concerns in children and adolescents, followed by child-friendly explanations from a child psychiatrist. Topics covered in these educational videos include: suicidal thoughts, lack of energy/motivation, anxiety, isolation and loneliness, obsessive worrying, attention difficulties, self-harm behaviors, sleep problems, and auditory hallucinations. The child psychiatrist appearing in these videos is one of the authors of this study (JF). The website also contains separate sections with mental health resources for children and families, including multiple Q&A entries about children’s mental health issues. These materials are purely informational and educational in nature, rather than providing interactive or personalized therapeutic interventions.

Unlike the intervention group, participants in the control group did not receive any structured therapeutic interaction through the AI chatbot. This design allowed for comparison between passive information provision (control) and active, personalized therapeutic engagement (intervention).

Participants in the control group underwent the same regular online evaluations and diary recording process as those in the intervention group.

### Criteria for Discontinuation

The criteria for discontinuing the intervention for individual participants were as follows:

1) Withdrawal of consent by the participant or their legal representative
2) Determination that the participant no longer met the inclusion criteria or met any exclusion criteria post-enrollment
3) Worsening of symptoms or findings that made study continuation challenging
4) Occurrence of adverse events that posed challenges to study continuation
5) Initiation of additional treatments, such as psychiatric care, counseling, or use of mental health applications
6) A determination by the principal investigator or sub-investigator that continuation was otherwise undesirable

The criteria for study termination were as follows:

1) Determination that the study intervention lacked expected efficacy, posed safety concerns, or was no longer meaningful to continue
2) Significant delays in case registration, frequent protocol deviations, or other factors that made study completion difficult
3) Occurrence of serious compliance issues affecting study execution.

### Primary Outcome

The primary outcome was the change in PHQ-9 scores from pre- to post-intervention. This measure was selected because assessing treatment efficacy through PHQ-9 score changes is commonly recommended and widely accepted in clinical research, including in Japan [20]. The PHQ-9, a self-administered questionnaire consisting of nine items, evaluates the presence and severity of depressive symptoms based on DSM-IV criteria for major depressive disorder within the past two weeks. The total PHQ-9 score ranges from 0 to 27, with higher scores indicating more severe depressive symptoms. The PHQ-9 score obtained at Week 0 was used as the baseline.

### Secondary Outcomes

The secondary outcome measures evaluated the correlation and relationship between changes in the primary outcome measure (PHQ-9) and the following items:

#### AIS

The AIS is a self-assessment tool developed as part of the World Health Organization’s (WHO) "World Project on Sleep and Health" to evaluate insomnia with high reliability and validity [21]. The scale includes eight items, with five assessing nighttime sleep difficulties ("sleep onset," "nighttime awakenings," "early morning awakenings," "sufficiency of total sleep duration," and "satisfaction with sleep quality") and three evaluating daytime functional impairment ("daytime mood," "daytime activity level," and "daytime sleepiness"). Participants rated the frequency of these experiences (at least three times a week in the past month) on a 4-point scale. A total score of 4 or more suggested suspected insomnia, while a score of 6 or more indicated insomnia.

#### Voice Analysis

During online consultations, participants’ voices were recorded while they conversed with the interviewer. Voice data was analyzed by SHIN4NY Inc., with recordings provided to the company for analysis. A previous study demonstrated that a speech emotion recognition model could predict depression [22].

#### Writing Pressure Analysis

Upon providing consent, participants received a diary and instructions on how to record entries using an intelligent pen pressure device developed by ZEBRA HOLDINGS Inc. This device, a mechanical pencil, automatically captured data on writing pressure, acceleration, and pen angle, storing it without requiring additional action from the participant. Previous research has suggested that an intelligent pen capable of measuring writing pressure may predict anxiety levels [23].

These data were collected at baseline (Week 0), during the intervention, and at the study’s conclusion (Week 9) through online consultations. Evaluations included PHQ-9, AIS, voice analysis, and intelligent pen pressure device data.

### Data Management

Data entry was conducted by research assistants and verified by the principal investigator. A data dictionary guided the coding process, and central monitoring took place once during the study period based on the collected case report forms.

This study managed data for both clinical outcomes and user engagement. In addition to the primary and secondary outcome measures, engagement metrics were tracked using CSV data logs provided by Emol Inc., including: 1) Total usage time, 2) Average daily usage time, 3) Usage time periods, 4) Last completed session, 5) Last usage date, 6) First usage date, 7) Session progression history. The purpose of analyzing these engagement metrics was twofold. First, we aimed to identify usage patterns and their potential relationship with changes in depressive symptoms. Second, these data were intended to assess whether participants engaged appropriately with the application and to evaluate whether they met the expected level of engagement for effective intervention. For example, adequate engagement might be defined as completing a minimum number of chatbot sessions or maintaining consistent interaction over time.

### Statistical Analysis

The analysis populations were defined as follows, with the primary analysis population being the Full Analysis Set (FAS). This study included all participants who were registered, randomized, received at least one session of the trial intervention, and had available efficacy data. Participants were excluded if baseline data were not obtained or if significant protocol violations occurred. Additionally, the FAS included participants who had no major deviations from the study protocol. Missing data were analyzed as observed without imputation, with the primary analysis performed on the full analysis set (FAS). As this study was exploratory in nature and lacked prior research in this specific population, no imputation for missing data was conducted.

The target sample size for the study was 60 participants (30 in each group). This calculation assumed an expected between-group mean difference in PHQ-9 change scores of 7, a standard deviation of 10, a two-sided significance level of 0.10, and a power of 0.80, using an independent two-sample t-test. The estimated sample size was 26 participants per group, and a target of 30 participants per group was set to allow for possible exclusions.

The primary analysis was conducted using the FAS. Changes in PHQ-9 scores from pre- to post-intervention were presented as the mean ± standard deviation. An analysis of covariance (ANCOVA) estimated the difference between groups, including the 90% confidence interval and p-value for the group comparison. A two-sided test was used, with statistical significance determined at P < 0.10. The secondary analysis was conducted on the Per Protocol Set (PPS) using the same summary and analytical methods as the primary analysis.

### Ethical Considerations

This study was registered with the Japan Registry of Clinical Trials (jRCT) under registration number jRCT1032230427. The full trial protocol is available on the jRCT website (Appendix). Approval for the study was granted by the Ethics Committee of Yokohama City University (Approval number F230907001). All procedures were conducted in accordance with the ethical standards outlined in the Declaration of Helsinki and the "Ethical Guidelines for Medical and Health Research Involving Human Subjects." Written informed consent was obtained from both participants and their guardians before enrollment. When possible, assent from participants was also sought; if direct assent was difficult to obtain, informed assent was acquired as an alternative. If participants exhibited severe psychological distress or suicidal ideation during the study, the research team had a protocol in place to guide them to appropriate psychiatric services. Emergency contact information for crisis intervention was provided to all participants and their guardians. The study adhered to safety monitoring procedures to ensure participant well-being throughout the intervention. Participant privacy and confidentiality were strictly protected, and data collected was used solely for research purposes. All personal information was handled in accordance with Yokohama City University’s Privacy Policy (Appendix). Findings from the study are to be disseminated through peer-reviewed journals and academic conferences.

### Protocol Version and Amendments

Protocol Version 1 was initially approved on September 27, 2023, with Version 2 subsequently approved on February 15, 2024.

## RESULTS

This study was conducted across four hospitals in Kanagawa Prefecture: Kanagawa Children’s Medical Center, Fujisawa City Hospital, Yokohama City University Hospital, and Yokohama City University Medical Center, targeting pediatric and adolescent psychiatric patients awaiting consultation. Recruitment materials were distributed from October 2023 to June 2024 for waiting list patients at each hospital as follows:

Kanagawa Children’s Medical Center: 78 patients
Fujisawa City Hospital: 8 patients
Yokohama City University Hospital: 10 patients
Yokohama City University Medical Center: 0 patients

A total of 96 patients received study invitations (78 from Kanagawa Children’s Medical Center, 8 from Fujisawa City Hospital, and 10 from Yokohama City University Hospital). Of these, 8 patients expressed interest by contacting us for additional information. Out of those who expressed interest, three patients scheduled and completed the informed consent (IC) process, while the remaining 5 did not proceed with IC due to lack of response when attempting to arrange a schedule.

Among the three patients who completed the informed consent process, one participant (a female adolescent) provided consent but subsequently withdrew from the study. The participant’s family initially contacted the research team on the scheduled day of the first online session, stating: "This morning, she became panic-stricken and is now unable to participate. Although it is the day of the appointment, would it be possible to cancel? I sincerely apologize for the inconvenience caused after all your preparations." In a follow-up message, the family elaborated: "She expressed anxiety about the online interview, making it impossible to proceed. We had hoped that engaging in this activity might help her develop a more positive outlook, but perhaps it was still too challenging for her." The other two patients who completed the informed consent process either declined participation due to concerns about diary recording requirements or were excluded after beginning medication at another facility. Another patient declined participation due to concerns about diary recording, and the third patient was excluded after beginning medication at another facility. Consequently, no evaluable data were obtained in this study.

## DISCUSSION

### Barriers to Engagement in Digital Interventions

The primary finding of this study - that children and adolescents with mental health challenges and their caregivers showed limited interest in an AI chatbot-based intervention while awaiting psychiatric consultation - highlights complex barriers to the adoption of digital mental health interventions. Several factors contributed to this low engagement. In this study, only one participant consented but subsequently withdrew due to a worsening of symptoms. This outcome suggests that patients with more severe symptoms may find digital interventions less suitable or accessible. However, given the need for timely intervention among high-severity cases, it is worth exploring ways to make AI-driven tools like chatbots more adaptable to their needs. Improving the usability and support level of digital tools for more severe cases could provide valuable early intervention options where in-person resources are limited.

### Digital Intervention Challenges: Accessibility, Family Influence, and Psychological Burden

First, one participant who withdrew provides valuable insights into digital intervention barriers. Despite digital tools being promoted as accessible for those with social anxiety, this case reveals that even virtual interactions can trigger significant psychological burden in adolescents with severe mental health challenges. The mother’s hope that "engaging in this activity might help her develop a more positive outlook" contrasted with her realization that "perhaps it was still too challenging for her," highlighting the gap between theoretical accessibility and practical barriers. The child psychiatrist who interviewed the patient and family during adverse event verification determined that the patient experienced increased subjective burden at the time of study participation, along with anxiety and avoidance symptoms exacerbated by her underlying depressive condition. This case demonstrates a "digital intervention paradox" - while technology aims to increase accessibility, implementation requirements like scheduled online sessions can create new barriers for the very individuals they intend to help. Future interventions might need truly asynchronous options that minimize direct interaction while maintaining efficacy and safety monitoring.

Many young patients, particularly those who have not received a formal diagnosis or treatment, may struggle to understand the relevance of app-based mental health support and may not fully appreciate its therapeutic value, especially when compared to the immediate effects associated with in-person interventions like pharmacotherapy or face-to-face therapy. A previous study with small sample sizes has shown that AI chatbot app dropouts for psychogenic premature ejaculation were only around 25% among Japanese adults [24]. On the other hand, previous studies suggest that young patients often lack intrinsic motivation to engage with mental health apps, especially when their symptoms are severe enough to require professional care [25,26]. In particular, another previous research indicates that individuals with more severe symptoms may benefit from more personalized support to enhance engagement and adherence to digital interventions [27]. Without a clear understanding of the intervention or trust in its benefits, these young patients may lack motivation to use the app consistently, contributing to low engagement in the study.

### Study Protocol Complexity and Privacy Concerns

This study may have unintentionally targeted a population less receptive to alternative digital interventions. Families who had already secured an upcoming psychiatric appointment may have seen little value in participating in a study involving digital interventions, preferring instead to wait for their scheduled in-person consultation. For these families, traditional in-person care may have appeared more reassuring, especially given the severity of the patient’s symptoms. Previous research on social influences in mental health service-seeking behavior among young people suggests that family is often the primary influence in choosing in-person services, whereas young people themselves tend to make decisions regarding online services [4]. Another study has also found that parents often seek informal support for their children’s mental health concerns initially, only turning to professional services as issues become more severe [28]. Additionally, patients with severe symptoms or their families often prefer in-person consultations over digital interventions, perceiving in-person care as more reliable and suitable for managing serious symptoms [29]. Therefore, patients and families may value the familiarity and perceived efficacy of traditional, in-person care as a more reliable or reassuring option compared to digital alternatives. This preference likely contributed to the reluctance toward digital solutions observed in this study. Engaging patients and families earlier in the mental health care process—before they have secured traditional clinical appointments—might improve receptiveness to digital options.

Furthermore, our study protocol involved multiple evaluation sessions, which could have imposed additional stress on participants, particularly those with social anxiety or other issues related to interpersonal interactions. Previous studies indicate that such requirements can increase dropout rates, especially among adolescents with social phobia [30,31]. In this context, the protocol’s demands may have discouraged participation and contributed to low engagement. Although efforts were made to reduce participant burden by conducting interviews online, the process of obtaining informed consent and providing detailed study information likely remained a challenge for some participants, especially those who may be sensitive to social interactions. Moreover, secondary assessment methods like voice and writing pressure analysis may have caused some participants to feel self-conscious or concerned about privacy, potentially exacerbating symptoms or causing reluctance to participate [32]. Privacy concerns are common with mental health apps, especially when personal data is analyzed, and this may affect user engagement [33]. In this study, participants may have felt uneasy about the voice recordings or writing pressure data being analyzed, as well as discussing their mental health status with researchers. These issues may have created additional barriers to engagement, particularly for young people unfamiliar with research environments. In the current study design, the exclusion of severe cases and the online interview assessment to reduce the burden of face-to-face implementation may not have been sufficient.

### Strengths and Limitations

This study had both methodological strengths and limitations. Conducting a randomized controlled trial in an understudied population—children and adolescents on psychiatric waiting lists—provided valuable insights into a critical phase of mental health service delivery, and the exploratory nature of the trial allowed for close examination of data acquisition and dropout rates. Additionally, our use of multiple assessment methods enabled a comprehensive evaluation of potential therapeutic effects and engagement patterns, which is rare in digital mental health studies involving youth.

However, the intensity of the intervention, including frequent evaluations and a structured protocol, may have been overwhelming for participants. This study suggests that less rigid protocols with fewer demands could enhance engagement. Additionally, recruiting patients already awaiting traditional psychiatric care may have reduced openness to digital alternatives, thus limiting the generalizability of our findings. Targeting a population that has not yet committed to in-person care—such as students in school counseling or those referred by community organizations—may address this issue in future studies. Finally, while secondary measures such as voice and writing pressure analysis provide valuable data, they may also create privacy concerns that deter participation, particularly among younger users. Additionally, typical limitations of eHealth trials should be noted. Participants were not blinded, which may have introduced performance bias. The informed consent process could have influenced their expectations, and the multiple outcomes planned for the study could have increased the risk of Type I errors. Future studies should address these biases and explore strategies to improve participant engagement. Furthermore, a key limitation of this study was the lack of a structured plan for conducting qualitative interviews and analyses on dropout reasons. This was primarily because the initial study design focused on quantitative outcome measures, and the feasibility of integrating additional qualitative assessments was not fully considered. While we identified some possible factors, such as social anxiety, depressive symptoms, and difficulties with maintaining engagement, a more systematic approach to understanding participants’ experiences and challenges would have provided deeper insights.

Future studies should incorporate structured qualitative methods, such as exit interviews or surveys, to better understand engagement barriers and develop targeted strategies for improving retention and adherence in digital mental health interventions for adolescents on psychiatric waiting lists. Previous qualitative studies have identified key factors influencing user engagement, including personalization, trust in AI, and perceived relevance of content [34]. Integrating these insights into future chatbot interventions may enhance usability and acceptability.

### Conclusions

To better understand the potential of AI chatbot interventions like “emol,” future studies could test the app in environments where supportive, pre-existing relationships exist, such as schools or community youth centers. Conducting trials in these familiar settings may foster trust, encourage participation, and enhance data validity, ultimately increasing the accessibility and effectiveness of digital mental health interventions for young people.

## Data Availability

All data produced in the present study are available upon reasonable request to the authors

## Acknowledgments

We would like to express our gratitude to the following individuals for their invaluable support in this study. Minori Ito assisted with participant recruitment and study-progress management as research assistant. Mikiko Yuzawa from Nouvelle Plus Inc. managed the data center and prepared the ethics application documents. Yasuyuki Okumura from Initiative for Clinical Epidemiological Research provided support in developing the research plan. We also thank Daiki Takekawa from Emol Inc., Eiji Okuno from ZEBRA HOLDINGS Inc., and Kanji Okazaki from SHIN4NY Inc. for their contributions in providing devices and support from their respective companies. We also acknowledge the assistance of OpenAI’s ChatGPT and Anthropic’s Claude in supporting the drafting and English editing process of this manuscript.

## Author Contributions

Conceived and designed the analysis: JF, SS, YY, AS, MT, TM, MI and TM conceived the study design and developed the analytical framework.

Data collection: JF, NS, MO, MT, and TM collected and curated the data used in the study.

Contributed data/analysis tools: JF, SS, MT, and TM provided essential data and developed analysis tools for the research.

Performed the analysis: JF, YY, SS conducted the data analysis and interpreted the results.

Wrote the paper: JF, YY, SS, MI, HY, MT, TM and MT drafted and revised the manuscript.

## Data Availability

The datasets generated and/or analyzed during the current study are not publicly available due to privacy protection and the scope of the research plan but are available from the corresponding author on reasonable request.

## Appendix List of Online Resources

- Yokohama City University child psychiatry department’s website http://www-user.yokohama-cu.ac.jp/~ycucap/
- Emol Inc. website https://emol.jp/
- jRCT website https://jrct.niph.go.jp/re/reports/detail/86750
- Yokohama City University’s Privacy Policy https://www.yokohama-cu.ac.jp/policy/privacy.html
- Report on age-specific population statistics of Kanagawa Prefecture https://www.pref.kanagawa.jp/docs/x6z/tc30/jinko/nenreibetu.html

## Supplementary Table

### Session structure and content of AI Chatbot emol based on acceptance and commitment therapy (ACT)

Each session integrated various content types including videos for psychological education and guided exercises, comics to illustrate metaphors, written practices, and conversation-based interactions. Content was designed to deliver ACT processes through multiple modalities: psychological education (introducing ACT concepts), metaphors (illustrating ACT principles through relatable stories and images that make abstract concepts concrete and accessible for children and adolescents), exercises (experiential practice of ACT skills), and practical applications (implementing ACT in daily life). Metaphors such as "feeding the anxiety trap," "tug of war with a monster," and "bus driver" were presented as comics to help young users intuitively understand complex psychological principles.

## Supplementary Figure

### Initial interaction with AI character "Roku"

The supplementary figure has been created to illustrate the initial user interaction with "Roku," the AI character in the emol application. The figure is organized into three panels (a, b, c) that demonstrate how the application establishes a supportive relationship through a conversational design that introduces users to the program structure and ACT framework.

#### Panel a: Introduction

This panel shows Roku introducing itself and the program. The conversation begins with a friendly greeting where Roku says "I’m Roku, and I’ll be supporting you throughout this program!" The panel then shows how Roku explains that the program is based on ACT (Acceptance and Commitment Therapy) and incorporates mindfulness techniques to help users deal with anxiety.

#### Panel b: Program Structure and ACT Framework

In this panel, Roku explains the commitment required ("It might be a bit challenging because nothing can be mastered without practice, but let’s do our best!") and introduces the concept of ACT. The character explains that ACT stands for Acceptance & Commitment Therapy, which is an approach to mental care by developing psychological flexibility to help manage anxiety and negative thoughts.

#### Panel c: Video-Based Learning Introduction

The final panel shows how the program transitions into structured learning content through video-based education. Roku introduces a mindfulness exercise video and prompts the user to watch it, with a "Have you finished watching?" question at the bottom to encourage engagement and interaction.

The interface uses simple, friendly language and visual elements specifically designed to engage adolescents. This introduction sets the foundation for the subsequent eight therapeutic sessions that progressively build psychological flexibility through ACT processes.

