## Supplementary table for "Challenges in Implementing a Mobile AI Chatbot Intervention for Depression Among Youth on Psychiatric Waiting Lists: A Randomized Control Study Termination Report"

| Session | Title | Summary of Contents | Duration |
|---|---|---|---|
| Session 1 | Distress and Suffering | • Introduction to ACT<br>• Role of anxiety<br>• Examining attempts to eliminate anxiety<br>• Explanation of awareness exercises | 15 min |
| Review 1 | Session 1 Review Session | Review of session 1 content | 8 min |
| Session 2 | Experiential Avoidance | • Examining avoidance patterns<br>• Recognizing the costs of avoidance<br>• Creative hopelessness<br>• "Feeding the anxiety trap" metaphor<br>• Observing anxiety rather than struggling with it<br>• Homework: "Things I've given up due to anxiety" | 18 min |
| Review 2 | Session 2 Review Session | Review of session 2 content | 12 min |
| Session 3 | What Can Be Controlled | • Control as the problem, not the solution<br>• "Tug of war with monster" metaphor<br>• Distinguishing between controllable and uncontrollable aspects<br>• Considering value-aligned actions<br>• Homework explanation | 15 min |
| Review 3 | Session 3 Review Session | Review of session 3 content | 10 min |
| Session 4 | Acceptance | • Acceptance and willingness<br>• Methods for facing uncomfortable emotions<br>• Exercise for accepting uncomfortable emotions | 18 min |
| Review 4 | Session 4 Review Session | Review of session 4 content | 13 min |
| Session 5 | Observer Perspective | • Volleyball metaphor for thoughts and feelings about anxiety | 10 min |
| Review 5 | Session 5 Review Session | Review of session 5 content | 5 min |
| Session 6 | Life Values | • Creating a values worksheet<br>• Life compass | 10-20 min |
| Review 6 | Session 6 Review Session | Review of session 6 content | 5-15 min |

| Session | Title | Summary of Contents | Duration |
| --- | --- | --- | --- |
| Session 7 | Commitment | • Willingness<br>• Creating a goal achievement table | 10-20 min |
| Review 7 | Session 7 Review Session | Review of session 7 content | 5-15 min |
| Session 8 | Continuing Commitment | • Activity verification<br>• Strategies for when things don't go well<br>• Summary | 5-15 min |
| Daily Training | Daily Training) | Same content as Session 7 review | 5-15 min |
| Reflection Session | Reflection Session | Same content as Session 8 "Continuing Commitment" | 5-15 min |
