## Supplementary figures and images for "Challenges in Implementing a Mobile AI Chatbot Intervention for Depression Among Youth on Psychiatric Waiting Lists: A Randomized Control Study Termination Report"

### Supplementary figure

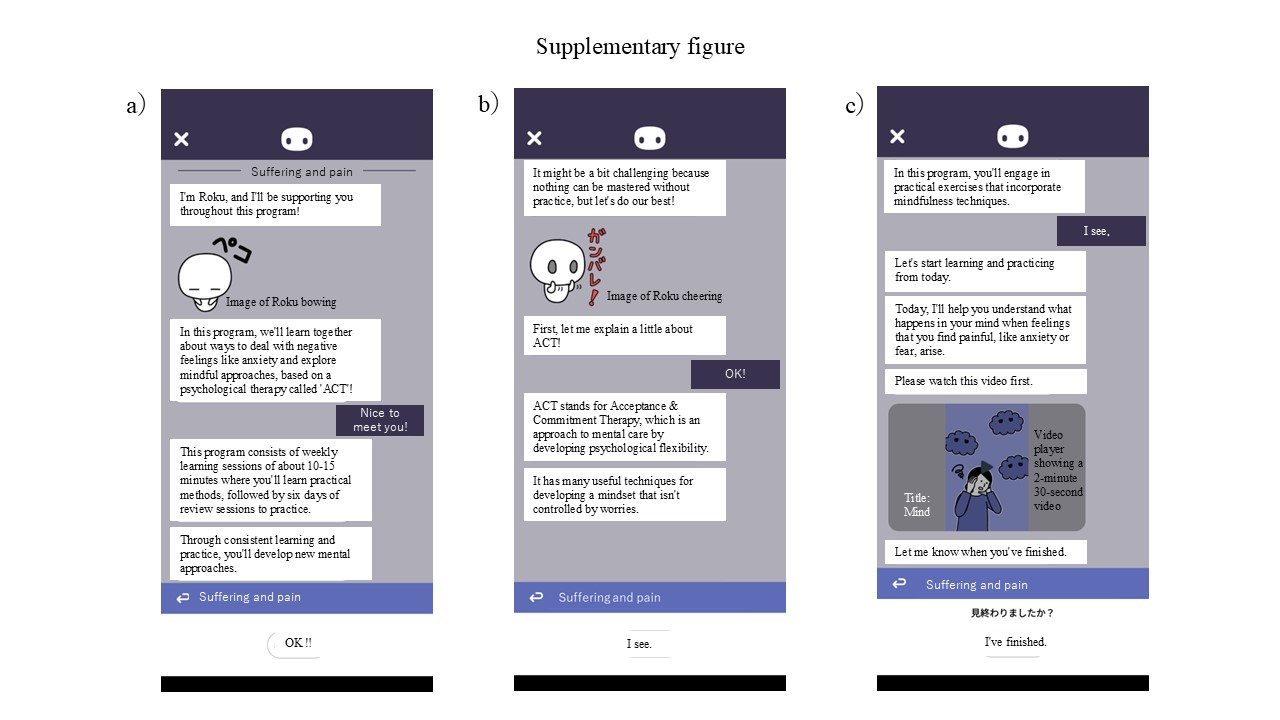
